# Low Blood Levels of Selenium, Selenoprotein P and GPx3 are Associated with Accelerated Biological Aging: Results from the Berlin Aging Study II (BASE-II)

**DOI:** 10.1101/2024.04.04.24305314

**Authors:** Valentin Max Vetter, Kamil Demircan, Jan Homann, Thilo Samson Chillon, Michael Mülleder, Orr Shomroni, Elisabeth Steinhagen-Thiessen, Markus Ralser, Christina M. Lill, Lars Bertram, Lutz Schomburg, Ilja Demuth

## Abstract

**Introduction:** Biological age reflects inter-individual differences in biological function and capacity beyond chronological age. Biological age can be estimated by DNA methylation age (DNAmA) and its deviation from chronological age, DNAmA acceleration (DNAmAA). Low levels of serum selenium, selenoprotein P (SELENOP), and the selenocysteine-containing glutathione peroxidase 3 (GPx3) are associated with adverse health outcomes and selenium supplementation is discussed as an anti-aging intervention.

**Methods:** In this study we analyzed 1,568 older participants from the Berlin Aging Study II (mean age +/− SD: 68.8 +/− 3.7 years, 51% women). DNAmA was estimated from genome-wide DNA methylation data using the Horvath, GrimAge, and DunedinPACE algorithms. Serum selenium levels were measured by total reflection X-ray fluorescence (TXRF) spectroscopy. SELENOP was measured by ELISA and GPx3 was derived from a larger set of mass spectrometry proteomics data.

**Results:** Participants with deficient serum selenium levels (<90μg/L) had a higher rate of biological aging (DunedinPACE, p=0.01, n=865). This association remained statistically significant after adjustment for age, sex, BMI, smoking, and genetic ancestry (β=-0.02, SE=0.01, 95%CI: - 0.034 to −0.004, n=757). Compared to the highest quartile, participants in the lowest quartile of SELENOP levels showed an accelerated biological aging rate (DunedinPACE, β=-0.03, SE=0.01, 95%CI: −0.051 to −0.008, n=740, fully adjusted model). Similarly, after adjustment for covariates, accelerated biological age was found in participants within the lowest GPx3 quartile compared to participants in the fourth quartile (DunedinPACE, p=<0.001 and GrimAge, p<0.001).

**Conclusion:** Our study suggests that low levels of selenium biomarkers are associated with accelerated biological aging measured as DNAmA. This effect was not substantially changed after adjustment for known covariates.

## Introduction

Over the past few decades the average lifespan increased faster than the healthspan, which resulted in a growing burden of late-life diseases (1, 2). However, there are substantial inter-individual differences in the pace of aging and some people remain disease-free and in good health until old age. Although some lifestyle habits were shown to impact some areas of aging (3) they do not fully explain the observed between-person differences. Hence, the identification of additional factors promoting a “healthy aging” trajectory would potentially benefit many. One recent, and often considered very useful (4, 5), biomarker to quantify difference in biological aging is DNA methylation age (DNAmA, epigenetic age) estimated from DNA methylation data by so called epigenetic clocks (6). The difference between epigenetic and chronological age, DNAmA acceleration (DNAmAA), was shown in previous studies to be associated with mortality as well as with numerous age-related diseases and phenotypes (6). Due to the ability of these clocks to assess the biological age of an individual they are increasingly used as endpoints for the evaluation of anti-aging interventions (reviewed in (7–11)).

In related research, one factor discussed in the context of healthy aging is selenium, an essential trace element that regulates thyroid hormone metabolism, antioxidative, and redox processes through incorporation into selenoproteins (12), which needs to be present in sufficient levels to exert its physiological function. Selenium status and its cellular availability (determined by dietary intake and its absorption) directly regulate expression and activity of selenoproteins (13). Selenium status can be determined by assessment of serum selenium biomarkers such as total selenium, the selenium transporter selenoprotein P (SELENOP), or glutathione peroxidase 3 (GPx3), a selenocysteine-containing extracellular antioxidant enzyme (14). Emerging clinical evidence indicates an association between a deficient status of these selenium biomarkers and outcomes of mortality and chronic diseases of older age, i.e., measures of an increased pace of aging (15–19). The direct association between selenium biomarkers and epigenetic aging, however, remains understudied. The relationship between blood selenium levels and DNAmA was analyzed in two previous studies, which revealed promising links between selenium levels and biological aging (20, 21). However, both studies were of small sample size (n=93 (20) and n=276 (21)), and analyzed only total selenium, but none of the other selenium-related variables. In this study, we analyzed the association between biological aging as determined with three DNAmA estimators (i.e. Horvath (22), GrimAge (23), and DunedinPACE (24)) total serum selenium, and two complementary protein biomarkers, SELENOP, GPx3, in a large cohort of 1,568 older participants from the Berlin Aging Study II (BASE-II).

## Methods

### Study population

The Berlin Aging Study II (BASE-II) is a multi-disciplinary study that aims to identify factors promoting healthy aging trajectories. BASE-II comprises 1,671 participants (60 – 85 years), who were recruited in the metropolitan area of Berlin, Germany from 2009 to 2014. Older participants who reported the intake of selenium supplements were excluded (n=103) resulting in a final dataset of up to 1,568 participants. A detailed description of the study’s cohort profile was published before and can be found in ref. (25).

### DNA methylation age (DNAmA), DNAmA acceleration (DNAmAA) and pace of aging

The participants’ epigenetic age was estimated from DNA methylation data utilizing three “first-generation” clocks (Horvath (22), Hannum (26), 7-CpG (27)), two “second-generation” clocks (PhenoAge (28), GrimAge (23)) and DunedinPACE (24), and a third-generation clock. DNAm data for the 7-CpG clock was measured by Single Nucleotide Prime Extension (SNuPE) in 1471 samples. The laboratory protocol is described in detail elsewhere (27). All other clocks were derived from DNAm data measured by the “Infinium MethylationEPIC” array (Illumina, Inc., USA), data of which was available for n=1,030 participants. Additional information on laboratory procedures and data processing can be found in reference (29). The DunedinPACE clock was calculated based using the methods described in the original publication (24).

DNA methylation age acceleration (DNAmAA) was calculated for all clocks (except for DunedinPACE) as residuals of a linear regression of DNAmA on chronological age accounting for leukocyte cell distribution (neutrophils, monocytes, lymphocytes, and eosinophils in G/l).

### Assessment of selenium, SELENOP and GPx3

The selenium biomarkers *SELENOP* and *GPx3* were measured at the Institute of Experimental Endocrinology, Charité University Hospital, Berlin, from serum samples that were taken during the same blood draw that produced the sample for the DNA isolation. Selenium status was measured by total reflection X-ray fluorescence (TXRF) spectroscopy as described in detail before (30, 31). SELENOP was measured from serum samples with a sandwich ELISA method using monoclonal human antibodies. Additional information on how selenium and SELENOP were measured in BASE-II were previously described in detail (32). GPx3 intensities were derived from proteomics data at baseline was measured by Liquid chromatography-mass spectrometry (LC-MS), using data independent acquisition (Supplementary Methods).

### Covariates

Chronological age, sex, and smoking behavior (in packyears) were assessed during a one-on-one interview with trained study personnel. Body weight and height were measured with the measuring station (763, SECA, Germany). Genetic ancestry was controlled for by using the first four principal components from a principal component analysis on genome-wide SNP genotyping data. A detailed description of the procedures applied to calculate this co-variate were published previously (33). Leukocyte cell distribution was measured from samples obtained at the same blood-draw that was used for DNA isolation in an accredited clinical biochemistry laboratory (MVZ Labor 28 GmbH, Berlin, Germany) by automated standard methods (flow cytometry).

### Statistical analysis

Statistical analyses were conducted in R (version 4.3.1) (34). Descriptive statistics were calculated with R’s tableone package (35). Linear regression analyses were calculated with R’s *lm* and *lm.beta* function (lm.beta package (36)) to examine the relationship between DNAmA estimators (dependent variable) and selenium biomarkers (independent variables). Figures were drawn with the ggplot2 package. Selenium biomarker outliers were defined as <25^th^ percentile −1.5*SD and >the 75^th^ percentile +1.5*SD. This resulted in the exclusion of n=46 (serum selenium), n=111 (SELENOP), and n=151 (GPx3) observations. Participants with missing values in phenotypic data were excluded from the respective analyses (available case analyses). Statistical significance was defined at α=5%. Due to the exploratory nature of this study, no adjustment was made for multiple testing.

## Results

### Study population and characteristics

The sample analyzed in this study comprised 1,568 BASE-II participants with a mean age of 69 years (SD=3.4, range 60-85 years). Sex was almost equally distributed (51% women). 656 (48.3%) participants had a selenium level of 90μg/L or lower, i.e. displayingselenium-deficiency. Study population characteristics are shown in Table 1.

**Table 1:** Baseline characteristics of analyzed sample.

| Variables | mean (SD), n (%) | Min | Max | n |
| --- | --- | --- | --- | --- |
| Age (years) | 68.79 (3.73) | 60,16 | 84,63 | 1568 |
| Sex (women) | 799 (51.0) |  |  | 1568 |
| BMI (kg/m<sup>2</sup>) | 26.83 (4.19) | 17,03 | 47,68 | 1534 |
| Smoking (packyears) | 10.51 (17.75) | 0,00 | 148,00 | 1511 |
| Serum Selenium (µg/L) | 89.65 (17.38) | 45,20 | 134,60 | 1357 |
| Selenium deficiency (<90 µg/L) | 656 (48.3) |  |  | 1357 |
| SELENOP (mg/L) | 3.72 (0.83) | 1,26 | 5,86 | 1324 |
| GPx3 (intensity) | 12.52 (0.27) | 11,33 | 13,11 | 1209 |
| 7-CpG DNAmAA (years) | 0.00 (6.97) | -22,93 | 26,61 | 1305 |
| Horvath DNAmAA (years) | 0.04 (4.27) | -14,71 | 17,90 | 853 |
| Hannum DNAmAA (years) | -0.04 (3.48) | -10,90 | 19,15 | 853 |
| PhenoAge DNAmAA (years) | 0.03 (4.61) | -16,88 | 14,28 | 853 |
| GrimAge DNAmAA (years) | 0.01 (3.08) | -8,52 | 10,20 | 853 |
| DunedinPACE (biol./chron.) | 1.01 (0.11) | 0,58 | 1,42 | 959 |

### Association of selenium with biological age acceleration

Visual inspections of the distribution of serum selenium levels vs. DNAmAA estimated by the GrimaAge and DunedinPACE clocks suggested possible non-linear associations (Figure 1). Therefore, in the next step we examined epigenetic age stratified by selenium deficiency (<90μg/L) and found a statistically significantly higher pace of aging measured by DunedinPACE in selenium-deficient participants (SMD=0.2, p=0.01, t-test, Figure 1 F). DunedinPACE remained higher in selenium-deficient participants after the adjustment for known covariates (chronological age, sex, BMI, smoking, genetic ancestry) in linear regression analyses (β=-0.02, SE=0.007, p=0.012, n=757, Table 2). Sex-stratified analyses showed that these results were mainly driven by the subgroup of men (Supplementary Table 2).

**Figure 1:**
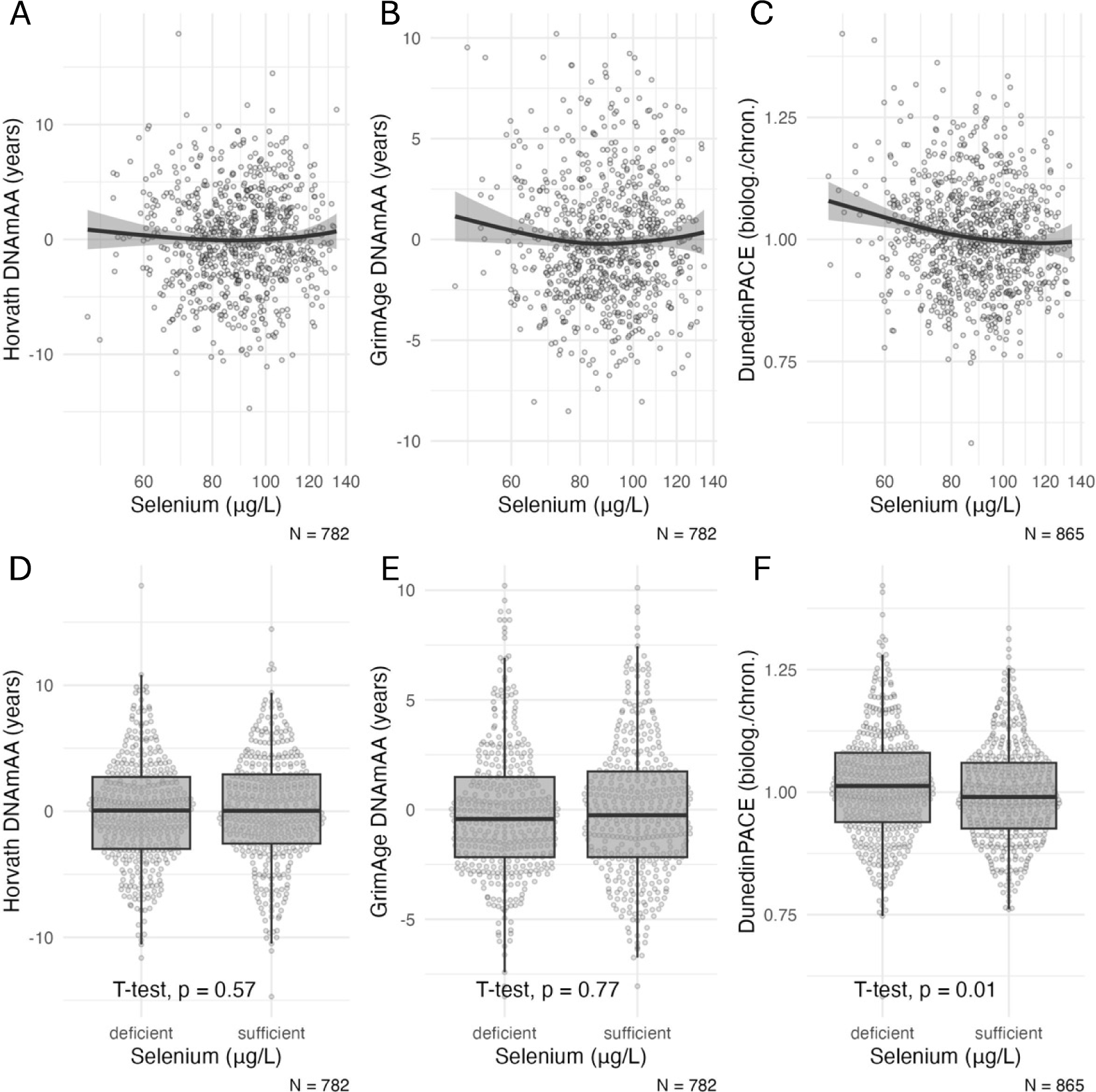
A-C: Scatterplots of serum selenium levels at baseline and biological age estimators. The x-axis is log-scaled. All available participants of the older age group are included. D-F: Boxplots of biological age estimators stratified by selenium status (deficient vs. sufficient. cut-off 90 μg/L) at baseline. Statistical significance of difference between group means was assessed by t-test. Note: DNAmAA = DNA methylation age acceleration; biolog. = biological years; chron. = chronological years. Boxplots show the median, hinges (corresponding to 25th and 75th percentile) and whiskers (1.5*inter-quartile range (IQR)).

**Table 2:**
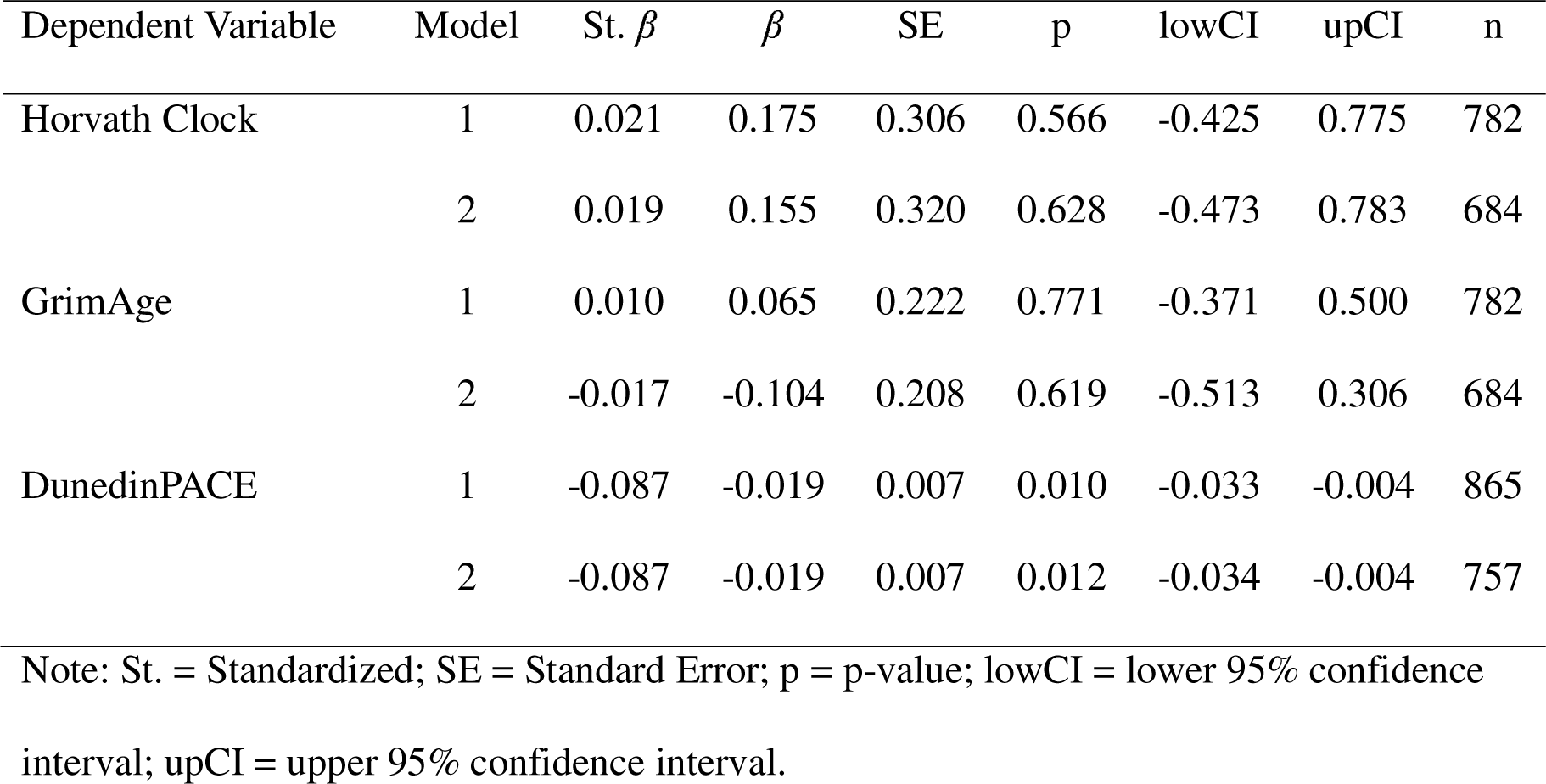
Linear regression analysis of epigenetic age estimators on selenium status (deficient vs. sufficient) at baseline. Model 1 is unadjusted. Model 2 is adjusted for chronological age, sex, BMI, smoking (packyears), and the first four genetic principal components (PC1 to PC4).

| Dependent Variable | Model | St. $\beta$ | $\beta$ | SE | p | lowCI | upCI | n |
| --- | --- | --- | --- | --- | --- | --- | --- | --- |
| Horvath Clock | 1 | 0.021 | 0.175 | 0.306 | 0.566 | -0.425 | 0.775 | 782 |
|  | 2 | 0.019 | 0.155 | 0.320 | 0.628 | -0.473 | 0.783 | 684 |
| GrimAge | 1 | 0.010 | 0.065 | 0.222 | 0.771 | -0.371 | 0.500 | 782 |
|  | 2 | -0.017 | -0.104 | 0.208 | 0.619 | -0.513 | 0.306 | 684 |
| DunedinPACE | 1 | -0.087 | -0.019 | 0.007 | 0.010 | -0.033 | -0.004 | 865 |
|  | 2 | -0.087 | -0.019 | 0.007 | 0.012 | -0.034 | -0.004 | 757 |

Note: St. = Standardized; SE = Standard Error; p = p-value; lowCI = lower 95% confidence interval; upCI = upper 95% confidence interval.

### Association of SELENOP with biological age

Low SELENOP levels were similarly associated with accelerated GrimAge DNAmAA and DunedinPACE (Figure 2). Assuming a non-linear relationship based on the scatterplot (Figure 2 A-C), SELENOP values were categorized into quartiles. A statistically significantly higher pace of biological aging (DunedinPACE) was measured in participants of the lowest quartile compared to participants in the highest quartile (SMD = 0.21, p=0.032, Figure 2 F). This association remained statistically significant after covariate adjustment (β=-0.03, SE=0.011, p=0.007, Table 3). After covariate adjustment the findings were only statistically significant in the subgroup of women (Supplementary Table 4 and 5).

**Figure 2:**
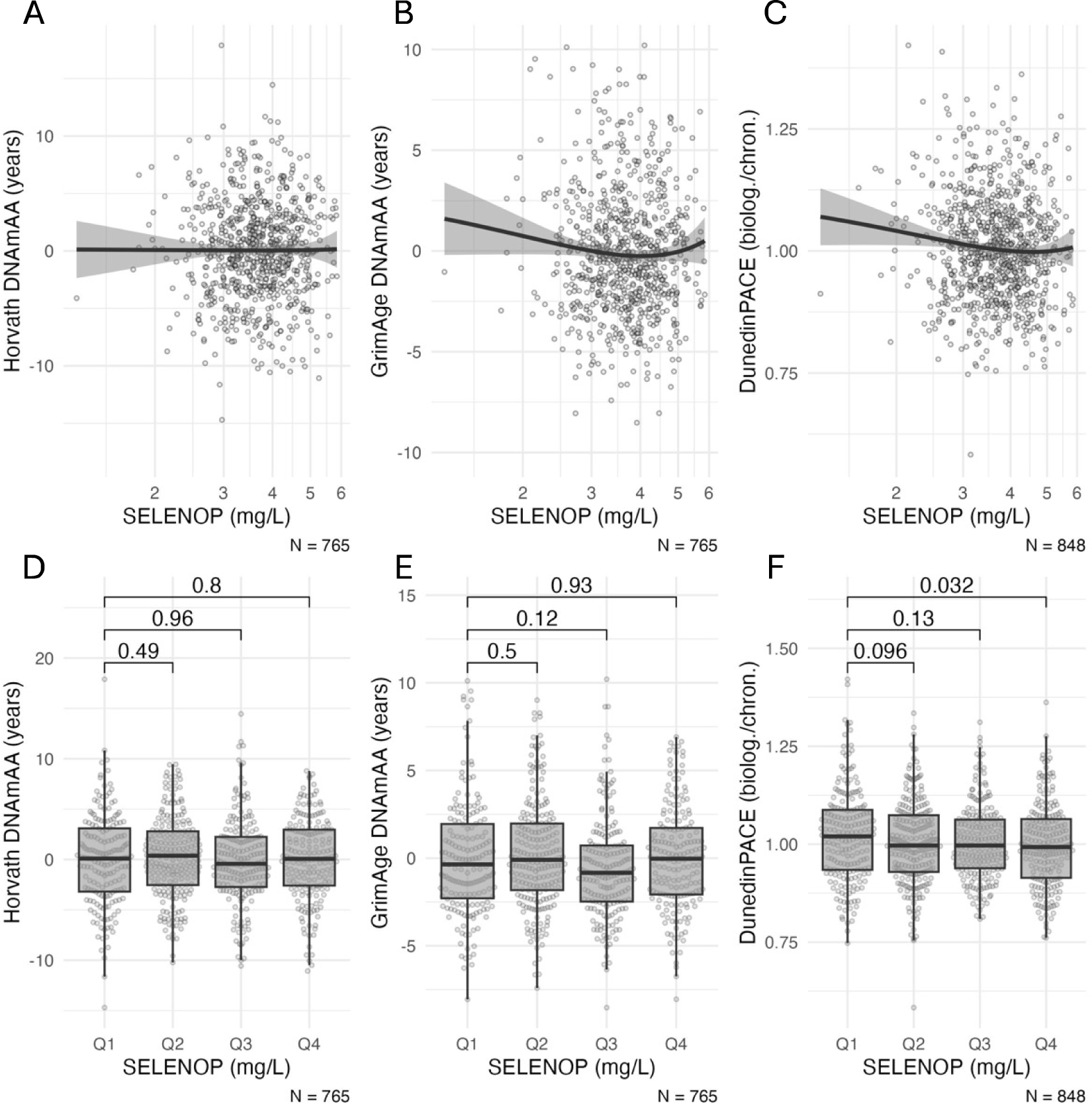
A-C: Scatterplots of SELENOP levels and biological age estimators. The x-axis is log-scaled. D-F: Boxplots of biological age estimators stratified by quartiles of SELENOP. Statistical significance of difference between group means was assessed by t-test. Note: DNAmAA = DNA methylation age acceleration; biolog. = biological years; chron. = chronological years. Boxplots show the median, hinges (corresponding to 25th and 75th percentile) and whiskers (1.5*inter-quartile range (IQR)).

**Table 3:**
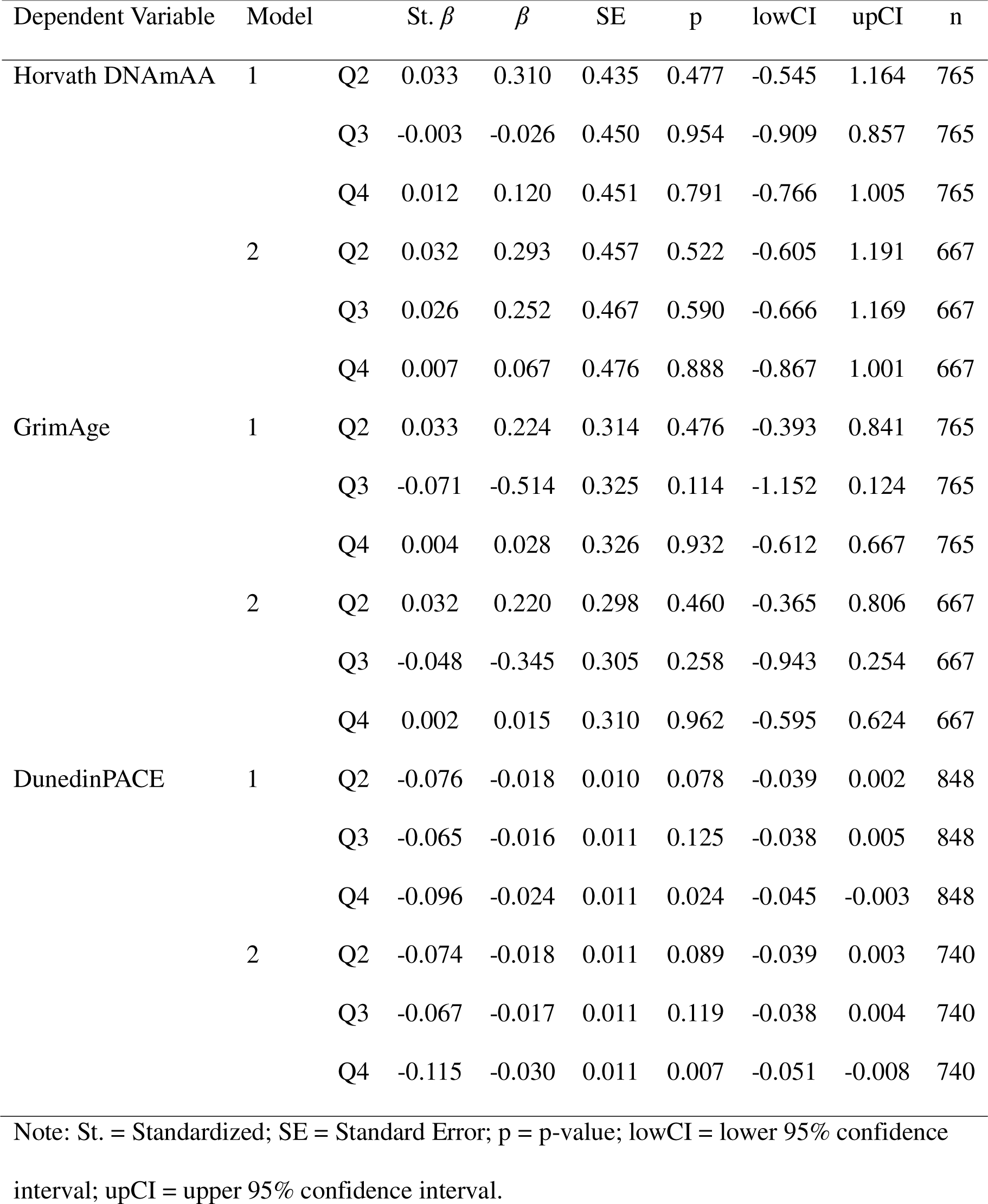
Linear regression analysis of epigenetic age estimators on quartiles of SELENOP at baseline. Model 1 is unadjusted. Model 2 is adjusted for chronological age, sex, BMI, smoking (packyears), and the first four genetic principal components (PC1 to PC4). The first quartile is used as reference.

### Association of GPx3 with biological age

Finally, compared to the lowest quartile, participants in the fourth quartile had a higher age acceleration estimated from the GrimAge clock (SMD = 0.41, p<0.001) and DunedinPACE clock (SMD = 0.54, p<0.001, Figure 3). These associations persisted to be statistically significant after covariate adjustment (p<=0.002, Table 4). Sex-specific analyses are presented in Supplementary Table 7 and 8.

**Figure 3:**
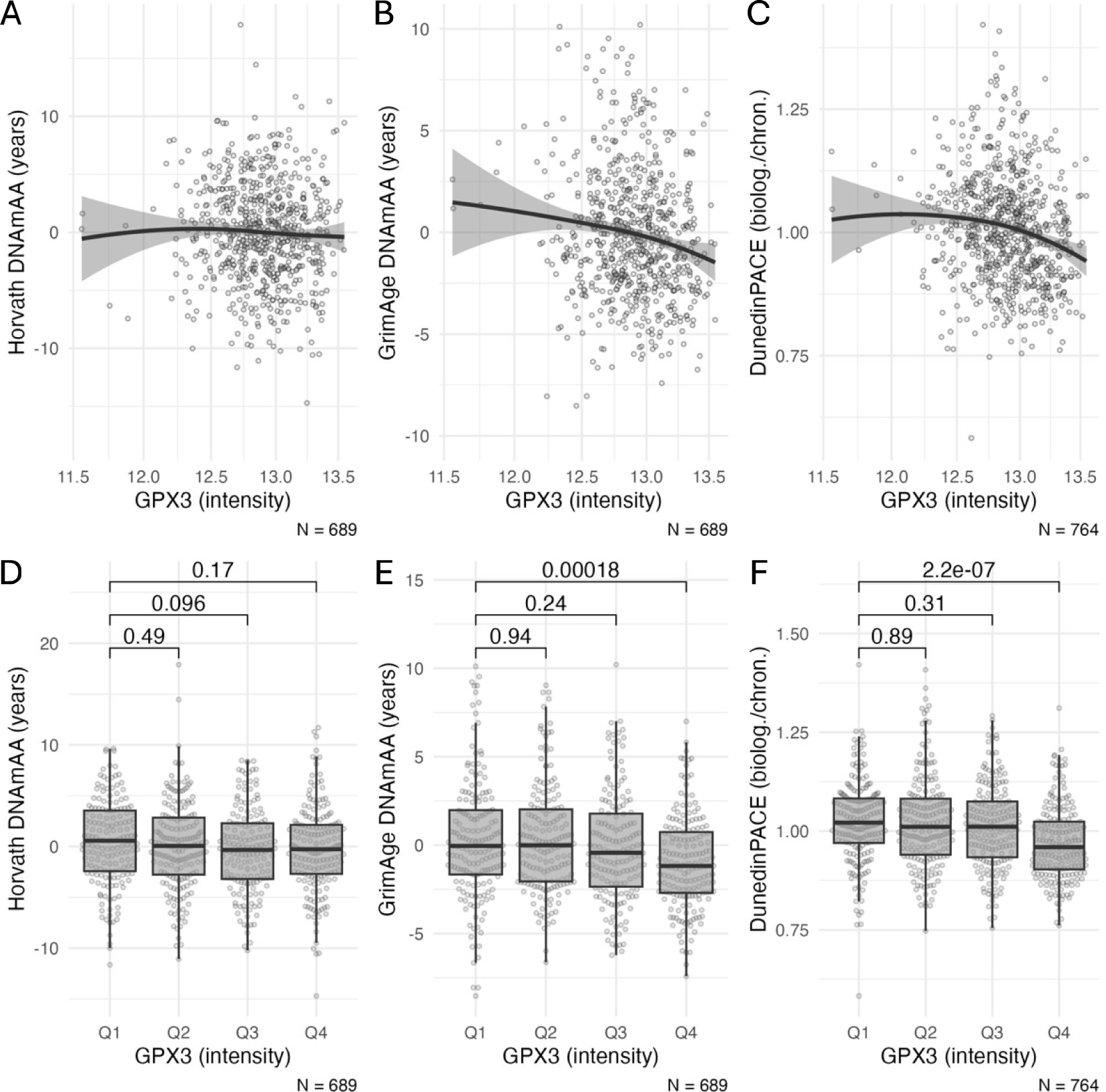
A-C: Scatterplots of GPx3 intensity (proteomics data) and biological age estimators. The x-axis is log-scaled. D-F: Boxplots of biological age estimators stratified by quartiles of GPx3 intensity (proteomics data). Statistical significance of difference between group means was assessed by t-test. Note: DNAmAA = DNA methylation age acceleration; biolog. = biological years; chron. = chronological years. Boxplots show the median, hinges (corresponding to 25th and 75th percentile) and whiskers (1.5*inter-quartile range (IQR)).

**Table 4:**
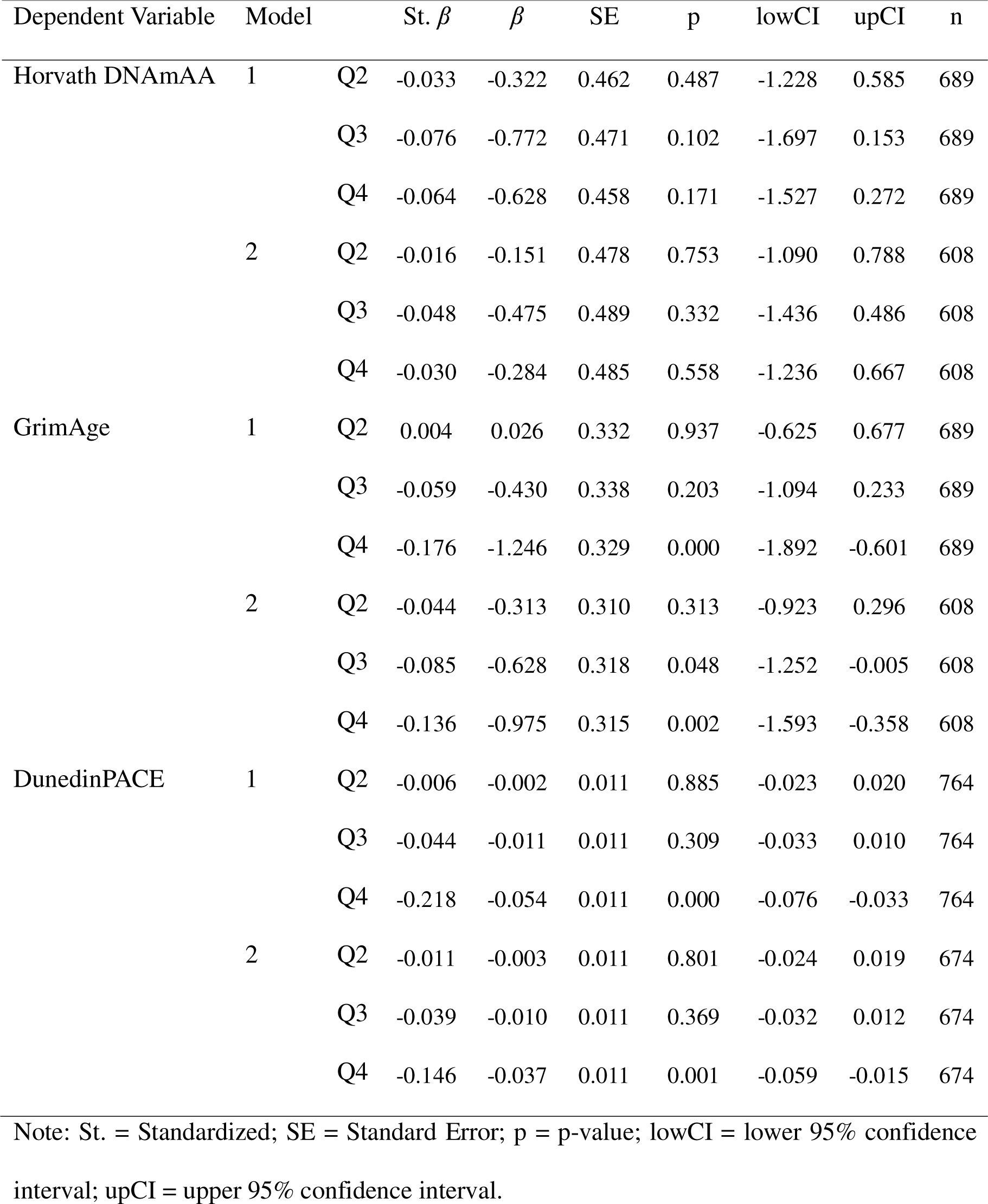
Linear regression analysis of epigenetic age estimators on quartiles of GPx3 intensity (proteomics data) at baseline. Model 1 is unadjusted. Model 2 is adjusted for chronological age, sex, BMI, smoking (packyears), and the first four genetic principal components (PC1 to PC4). The first quartile is used as reference group.

## Discussion

In this study, we analyzed the association between serum biomarkers, namely total serum selenium, SELENOP, GPx3 and biological age measured by epigenetic clocks in 865 participants of BASE-II. Lower values in all three selenium biomarkers were associated with an increased pace of aging measured with the DunedinPACE clock. This association was independent from covariates included in the linear regression analysis.

The stepwise increase in the strength of association from the first- to the third-generation clock is likely due to the methodological approach used for their development with the later clocks potentially including CpG sites that better reflect the underlying aging process (37). Furthermore, newer DNAmA clocks are more sensitive towards exogenous effects (20). This trend was also observed with clinical phenotypes (7, 24, 38).

Our findings are in line with results published by Cheng and colleagues (20) who found a statistically significant association between serum selenium levels and DunedinPACE in a sample of 93 participants. In contrast, we could not validate the association between selenium levels and PhenoAge DNAmAA reported by the same authors (Supplementary Table 2). Furthermore, regarding our non-significant associations of selenium levels with earlier DNAmA clocks, our results are consistent with findings from a cohort of 276 older Chinese men and women that showed no statistically significant association between selenium and the first- and second-generation clocks (21).

Boyer and colleagues did not find a consistent association between urine selenium and any epigenetic age estimation in linear regression models in an American Indian population of 2,301 participants (Strong Heart Study, SHS) (37). However, when analyzing the effect of the total metal mixture using Bayesian Kernel Machine Regression, a non-linear association with PhenoAge, GrimAge and DunedinPACE was found (37). The difference in these results compared to our study may arise from the choice of matrix used to determine selenium status. Specifically, the selenium status in the SHS was replete (37), whereas BASE-II participants showed a borderline selenium-deficiency. The lack of effects observed in SHS may also be attributable to the saturated expression of selenoproteins, and thus likely reflecting a threshold effect as commonly observed for selenium and health outcomes.

Our analyses do not allow to draw any conclusions on cause-effect relationships between selenium levels and accelerated biological aging. However, our results corroborate recent findings on aging phenotypes assessed by other clinical and phenotypical outcomes that show an association between selenium biomarkers and mortality (15, 19, 39), cardiovascular outcomes (16, 17, 39), and several cancer entities (14, 40–42). Moreover, an effect of selenium levels on epigenetic age appears biologically plausible since it is well established that selenium levels in rodents, cell-lines (human, mouse) and human tissue impact the methylome (reviewed in (43)).

Strengths of this study include the large sample size with an equal representation of men and women and a high degree of data completeness for the study parameters and potential confounders. Furthermore, this is the first study that examines additional selenium biomarkers, i.e., SELENOP and GPx3, in addition to selenium levels. The differing half-lives of these biomarkers ensure low risk of misclassification due to e.g., prior supplementation or enriched dietary selenium intake.

However, our study also has a few limitations. Firstly, as outlined above, the cross-sectional analyses presented in this study do not allow to draw any conclusions about causality or direction of effect. Secondly, the analyzed sample is of above-average health. Therefore, it is possible that the effects reported here are underestimated since BASE-II participants are generally healthier. In addition, this limits the generalizability of our results to the background population of mainly European elderly subjects with borderline sufficient selenium status residing in a metropolitan area. Thirdly, due to the explorative nature of this analysis, no adjustment for multiple testing was made. Fourthly, no detailed information about dosage and frequency of potential additional selenium intake by nutritional supplements was available. Therefore, all participants who reported selenium supplementation were excluded and evaluating effects of selenium supplementation on biological age was precluded.

## Conclusion

Our study shows that selenium biomarkers are moderately associated with accelerated biological aging in a European-based dataset. These results support previous findings that suggest health benefits of sufficient selenium levels. Randomized controlled trials are needed to evaluate the potential therapeutic benefit of selenium supplementation and its effect on biological age acceleration.

## Data Availability

Due to concerns for participant privacy, data are available only upon reasonable request. Please contact Ludmila MuCller, scientific coordinator, at, for additional information.

## Ethics

All participants gave written informed consent. The Ethics Committee of the Charité – Universitätsmedizin Berlin approved the study (approval numbers EA2/029/09 and EA2/144/16). The study was conducted in accordance with the Declaration of Helsinki and was registered in the German Clinical Trials Registry as DRKS00009277.

## Author contributions

Conceptualization: V.M.V., K.D., T.S.C.; L.S., I.D.; Data curation: V.M.V., J.H., M.M., O.S., L.B., I.D.; Formal analysis: V.M.V.; Funding acquisition: E.S.-T.; M.R., L.B., L.S., I.D.; Resources: E.S.-T.; M.R., L.B., L.S., I.D.; Supervision: C.L., M.R., L.B., L.S., I.D.; Visualization: V.M.V.; Writing: V.M.V, K.D., I.D.; Reviewed and approved the manuscript: all authors.

