## Supplementary Material for "Low Blood Levels of Selenium, Selenoprotein P and GPx3 are Associated with Accelerated Biological Aging: Results from the Berlin Aging Study II (BASE-II)"

Methods:

*Mesurement of GPx3*

The sample preparation involved semi-automated processing of serum samples in 96-well plates, including denaturation, reduction, alkylation, trypsin digestion, and cleanup steps (34). Liquid chromatography-mass spectrometry (LC-MS) analysis was conducted using a Bruker timsTOF Pro system coupled with an Agilent 1290 Infinity II LC system (35). Computational proteomics involved generating a spectral library based on Human Plasma PeptideAtlas and annotating peptide sequences to the Uniprot human reference proteome (36, 37). The software DIA-NN was used for data annotation and quantification (38). Pre-processing was performed using MS-DAP (39) as framework and included data normalization, outlier sample filtering, low-presence peptide filtering, data imputation, and batch correction to ensure high data quality.

Tables:

Supplementary Table 1: Standardized mean difference of epigenetic age estimators between selenium deficient participants and selenium sufficient participants. Statistical significance of difference was assessed by t-test.

| Variable | Deficient Mean(SD) | n | Sufficient Mean (SD) | n | p | SMD |
|---|---|---|---|---|---|---|
| Horvath DNAmAA | -0.03 (4.20) | 391 | 0.15 (4.34) | 391 | 0.566 | 0.04 |
| GrimAge DNAmAA | -0.11 (3.08) | 391 | -0.04 (3.13) | 391 | 0.771 | 0.02 |
| DunedinPACE | 1.02 (0.11) | 440 | 1.00 (0.10) | 425 | 0.010 | 0.18 |

Note: SD = Standard Deviation, n = number of observations, SMD = Standardized Mean Difference.

Supplementary Table 2: Linear regression analysis of epigenetic age estimators calculated from all six epigenetic clocks available on selenium status (deficient vs. sufficient) in the complete dataset as well as sex-stratified subgroups. Model 1 is unadjusted. Model 2 is adjusted for chronological age, sex, BMI, smoking (packyears), and the first four genetic principal components (PC1 to PC4).

| Clock | Model | St. $\beta$ | $\beta$ | SE | p | lowCI | upCI | n |
|---|---|---|---|---|---|---|---|---|
| Women and Men | | | | | | | | |
| 7-CpG DNAmAA | 1 | 0.022 | 0.298 | 0.396 | 0.452 | -0.480 | 1.075 | 1198 |
| | 2 | 0.004 | 0.058 | 0.521 | 0.912 | -0.965 | 1.081 | 691 |
| Horvath DNAmAA | 1 | 0.021 | 0.175 | 0.306 | 0.566 | -0.425 | 0.775 | 782 |
| | 2 | 0.019 | 0.155 | 0.320 | 0.628 | -0.473 | 0.783 | 684 |
| Hannum DNAmAA | 1 | 0.002 | 0.015 | 0.250 | 0.953 | -0.475 | 0.505 | 782 |
| | 2 | 0.002 | 0.015 | 0.262 | 0.955 | -0.499 | 0.529 | 684 |
| PhenoAge DNAmAA | 1 | -0.036 | -0.330 | 0.332 | 0.320 | -0.983 | 0.322 | 782 |
| | 2 | -0.038 | -0.344 | 0.352 | 0.328 | -1.034 | 0.346 | 684 |
| GrimAge DNAmAA | 1 | 0.010 | 0.065 | 0.222 | 0.771 | -0.371 | 0.500 | 782 |
| | 2 | -0.017 | -0.104 | 0.208 | 0.619 | -0.513 | 0.306 | 684 |
| DunedinPACE | 1 | -0.087 | -0.019 | 0.007 | 0.010 | -0.033 | -0.004 | 865 |
| | 2 | -0.087 | -0.019 | 0.007 | 0.012 | -0.034 | -0.004 | 757 |
| Women | | | | | | | | |
| 7-CpG DNAmAA | 1 | 0.054 | 0.713 | 0.542 | 0.189 | -0.352 | 1.778 | 602 |
| | 2 | 0.088 | 1.185 | 0.725 | 0.103 | -0.241 | 2.612 | 357 |
| Horvath DNAmAA | 1 | 0.025 | 0.205 | 0.415 | 0.621 | -0.611 | 1.021 | 400 |
| | 2 | 0.041 | 0.340 | 0.446 | 0.446 | -0.537 | 1.218 | 354 |
| Hannum DNAmAA | 1 | -0.024 | -0.155 | 0.327 | 0.637 | -0.798 | 0.489 | 400 |
| | 2 | 0.001 | 0.007 | 0.350 | 0.984 | -0.682 | 0.695 | 354 |
| PhenoAge DNAmAA | 1 | -0.051 | -0.476 | 0.468 | 0.310 | -1.396 | 0.444 | 400 |
| | 2 | -0.055 | -0.518 | 0.499 | 0.300 | -1.500 | 0.464 | 354 |
| GrimAge DNAmAA | 1 | 0.018 | 0.103 | 0.278 | 0.713 | -0.444 | 0.649 | 400 |
| | 2 | -0.005 | -0.027 | 0.288 | 0.924 | -0.594 | 0.539 | 354 |
| DunedinPACE | 1 | -0.083 | -0.017 | 0.010 | 0.081 | -0.035 | 0.002 | 447 |
| | 2 | -0.066 | -0.013 | 0.010 | 0.176 | -0.032 | 0.006 | 395 |
| Men | | | | | | | | |
| 7-CpG DNAmAA | 1 | -0.018 | -0.241 | 0.566 | 0.670 | -1.353 | 0.870 | 596 |
| | 2 | -0.078 | -1.062 | 0.752 | 0.159 | -2.542 | 0.418 | 334 |
| Horvath DNAmAA | 1 | 0.001 | 0.007 | 0.439 | 0.987 | -0.856 | 0.871 | 382 |
| | 2 | -0.009 | -0.077 | 0.465 | 0.869 | -0.992 | 0.838 | 330 |
| Hannum DNAmAA | 1 | 0.007 | 0.051 | 0.364 | 0.889 | -0.665 | 0.767 | 382 |
| | 2 | -0.006 | -0.044 | 0.396 | 0.912 | -0.823 | 0.735 | 330 |
| PhenoAge DNAmAA | 1 | -0.027 | -0.246 | 0.471 | 0.602 | -1.172 | 0.680 | 382 |
| | 2 | -0.022 | -0.195 | 0.500 | 0.697 | -1.178 | 0.788 | 330 |
| GrimAge DNAmAA | 1 | -0.031 | -0.184 | 0.309 | 0.552 | -0.793 | 0.424 | 382 |
| | 2 | -0.030 | -0.185 | 0.305 | 0.546 | -0.786 | 0.416 | 330 |
| DunedinPACE | 1 | -0.119 | -0.026 | 0.011 | 0.015 | -0.047 | -0.005 | 418 |
| | 2 | -0.109 | -0.024 | 0.011 | 0.034 | -0.047 | -0.002 | 362 |

Note: St. = Standardized; SE = Standard Error; p = p-value; lowCI = lower 95% confidence interval; upCI = upper 95% confidence interval.

Supplementary Table 3: Linear regression analysis of epigenetic age estimates calculated by all available six epigenetic clocks on quartiles of Selenoprotein P in men and women. Model 1 is unadjusted. Model 2 is adjusted for chronological age, sex, BMI, smoking (packyears), and the first four genetic principal components (PC1 to PC4). The first quartile is used as reference.

| Clock | Model | | St. $\beta$ | $\beta$ | SE | p | lowCI | upCI | n |
|---|---|---|---|---|---|---|---|---|---|
| 7-CpG DNAmAA | 1 | Q2 | -0.018 | -0.284 | 0.563 | 0.613 | -1.388 | 0.820 | 1168 |
| | | Q3 | -0.033 | -0.525 | 0.567 | 0.354 | -1.637 | 0.587 | 1168 |
| | | Q4 | 0.007 | 0.109 | 0.568 | 0.847 | -1.005 | 1.224 | 1168 |
| | 2 | Q2 | 0.051 | 0.771 | 0.734 | 0.294 | -0.671 | 2.213 | 674 |
| | | Q3 | -0.001 | -0.020 | 0.753 | 0.979 | -1.497 | 1.458 | 674 |
| | | Q4 | 0.022 | 0.348 | 0.764 | 0.649 | -1.152 | 1.848 | 674 |
| Horvath DNAmAA | 1 | Q2 | 0.033 | 0.310 | 0.435 | 0.477 | -0.545 | 1.164 | 765 |
| | | Q3 | -0.003 | -0.026 | 0.450 | 0.954 | -0.909 | 0.857 | 765 |
| | | Q4 | 0.012 | 0.120 | 0.451 | 0.791 | -0.766 | 1.005 | 765 |
| | 2 | Q2 | 0.032 | 0.293 | 0.457 | 0.522 | -0.605 | 1.191 | 667 |
| | | Q3 | 0.026 | 0.252 | 0.467 | 0.590 | -0.666 | 1.169 | 667 |
| | | Q4 | 0.007 | 0.067 | 0.476 | 0.888 | -0.867 | 1.001 | 667 |
| Hannum DNAmAA | 1 | Q2 | 0.111 | 0.857 | 0.354 | 0.016 | 0.162 | 1.551 | 765 |
| | | Q3 | -0.023 | -0.184 | 0.366 | 0.616 | -0.901 | 0.534 | 765 |
| | | Q4 | 0.004 | 0.031 | 0.367 | 0.932 | -0.688 | 0.751 | 765 |
| | 2 | Q2 | 0.132 | 1.002 | 0.372 | 0.007 | 0.273 | 1.732 | 667 |
| | | Q3 | 0.011 | 0.086 | 0.380 | 0.821 | -0.659 | 0.831 | 667 |
| | | Q4 | 0.012 | 0.098 | 0.386 | 0.801 | -0.661 | 0.857 | 667 |
| PhenoAge DNAmAA | 1 | Q2 | 0.024 | 0.252 | 0.475 | 0.596 | -0.681 | 1.184 | 765 |
| | | Q3 | -0.024 | -0.255 | 0.491 | 0.604 | -1.219 | 0.709 | 765 |
| | | Q4 | -0.016 | -0.177 | 0.492 | 0.719 | -1.144 | 0.790 | 765 |
| | 2 | Q2 | 0.026 | 0.269 | 0.505 | 0.594 | -0.722 | 1.261 | 667 |
| | | Q3 | -0.027 | -0.287 | 0.516 | 0.578 | -1.300 | 0.726 | 667 |
| | | Q4 | -0.039 | -0.417 | 0.525 | 0.427 | -1.449 | 0.614 | 667 |
| GrimAge DNAmAA | 1 | Q2 | 0.033 | 0.224 | 0.314 | 0.476 | -0.393 | 0.841 | 765 |
| | | Q3 | -0.071 | -0.514 | 0.325 | 0.114 | -1.152 | 0.124 | 765 |
| | | Q4 | 0.004 | 0.028 | 0.326 | 0.932 | -0.612 | 0.667 | 765 |
| | 2 | Q2 | 0.032 | 0.220 | 0.298 | 0.460 | -0.365 | 0.806 | 667 |
| | | Q3 | -0.048 | -0.345 | 0.305 | 0.258 | -0.943 | 0.254 | 667 |
| | | Q4 | 0.002 | 0.015 | 0.310 | 0.962 | -0.595 | 0.624 | 667 |
| DunedinPACE | 1 | Q2 | -0.076 | -0.018 | 0.010 | 0.078 | -0.039 | 0.002 | 848 |
| | | Q3 | -0.065 | -0.016 | 0.011 | 0.125 | -0.038 | 0.005 | 848 |
| | | Q4 | -0.096 | -0.024 | 0.011 | 0.024 | -0.045 | -0.003 | 848 |
| | 2 | Q2 | -0.074 | -0.018 | 0.011 | 0.089 | -0.039 | 0.003 | 740 |
| | | Q3 | -0.067 | -0.017 | 0.011 | 0.119 | -0.038 | 0.004 | 740 |
| | | Q4 | -0.115 | -0.030 | 0.011 | 0.007 | -0.051 | -0.008 | 740 |

Note: St. = Standardized; SE = Standard Error; p = p-value; lowCI = lower 95% confidence interval; upCI = upper 95% confidence interval.

Supplementary Table 4: Linear regression analysis of epigenetic age estimates calculated by all available six epigenetic clocks on quartiles of Selenoprotein P in the subgroup of women. Model 1 is unadjusted. Model 2 is adjusted for chronological age, sex, BMI, smoking (packyears), and the first four genetic principal components (PC1 to PC4). The first quartile is used as reference.

| Clock | Model | | St. $\beta$ | $\beta$ | SE | p | lowCI | upCI | n |
|---|---|---|---|---|---|---|---|---|---|
| 7-CpG DNAmAA | 1 | Q2 | 0.027 | 0.407 | 0.760 | 0.593 | -1.085 | 1.898 | 588 |
| | | Q3 | 0.018 | 0.265 | 0.764 | 0.729 | -1.236 | 1.766 | 588 |
| | | Q4 | 0.084 | 1.272 | 0.772 | 0.100 | -0.245 | 2.789 | 588 |
| | 2 | Q2 | 0.126 | 1.815 | 1.002 | 0.071 | -0.156 | 3.785 | 348 |
| | | Q3 | 0.025 | 0.365 | 1.015 | 0.719 | -1.631 | 2.361 | 348 |
| | | Q4 | 0.044 | 0.688 | 1.069 | 0.520 | -1.414 | 2.791 | 348 |
| Horvath DNAmAA | 1 | Q2 | 0.096 | 0.877 | 0.589 | 0.137 | -0.281 | 2.036 | 391 |
| | | Q3 | 0.083 | 0.786 | 0.603 | 0.194 | -0.400 | 1.972 | 391 |
| | | Q4 | 0.061 | 0.603 | 0.622 | 0.333 | -0.620 | 1.826 | 391 |
| | 2 | Q2 | 0.045 | 0.412 | 0.633 | 0.516 | -0.833 | 1.657 | 345 |
| | | Q3 | 0.072 | 0.669 | 0.638 | 0.295 | -0.586 | 1.925 | 345 |
| | | Q4 | 0.035 | 0.345 | 0.675 | 0.609 | -0.982 | 1.673 | 345 |
| Hannum DNAmAA | 1 | Q2 | 0.137 | 0.993 | 0.463 | 0.033 | 0.083 | 1.904 | 391 |
| | | Q3 | 0.036 | 0.271 | 0.474 | 0.568 | -0.661 | 1.203 | 391 |
| | | Q4 | 0.021 | 0.165 | 0.489 | 0.736 | -0.796 | 1.126 | 391 |
| | 2 | Q2 | 0.163 | 1.162 | 0.493 | 0.019 | 0.192 | 2.131 | 345 |
| | | Q3 | 0.050 | 0.362 | 0.497 | 0.466 | -0.615 | 1.340 | 345 |
| | | Q4 | 0.025 | 0.192 | 0.525 | 0.716 | -0.842 | 1.225 | 345 |
| PhenoAge DNAmAA | 1 | Q2 | 0.075 | 0.772 | 0.665 | 0.247 | -0.536 | 2.080 | 391 |
| | | Q3 | 0.006 | 0.069 | 0.681 | 0.920 | -1.271 | 1.408 | 391 |
| | | Q4 | 0.037 | 0.418 | 0.703 | 0.552 | -0.964 | 1.799 | 391 |
| | 2 | Q2 | 0.055 | 0.567 | 0.709 | 0.424 | -0.827 | 1.962 | 345 |
| | | Q3 | -0.005 | -0.053 | 0.715 | 0.941 | -1.459 | 1.353 | 345 |
| | | Q4 | 0.011 | 0.129 | 0.756 | 0.865 | -1.358 | 1.616 | 345 |
| GrimAge DNAmAA | 1 | Q2 | 0.054 | 0.329 | 0.394 | 0.405 | -0.446 | 1.103 | 391 |
| | | Q3 | -0.068 | -0.433 | 0.403 | 0.284 | -1.226 | 0.360 | 391 |
| | | Q4 | -0.001 | -0.009 | 0.416 | 0.984 | -0.826 | 0.809 | 391 |
| | 2 | Q2 | 0.051 | 0.319 | 0.407 | 0.434 | -0.482 | 1.120 | 345 |
| | | Q3 | -0.079 | -0.505 | 0.411 | 0.219 | -1.313 | 0.303 | 345 |
| | | Q4 | -0.016 | -0.112 | 0.434 | 0.796 | -0.966 | 0.742 | 345 |
| DunedinPACE | 1 | Q2 | -0.101 | -0.023 | 0.014 | 0.095 | -0.049 | 0.004 | 438 |
| | | Q3 | -0.058 | -0.013 | 0.014 | 0.335 | -0.040 | 0.014 | 438 |
| | | Q4 | -0.108 | -0.026 | 0.014 | 0.068 | -0.054 | 0.002 | 438 |
| | 2 | Q2 | -0.086 | -0.019 | 0.014 | 0.161 | -0.046 | 0.008 | 386 |
| | | Q3 | -0.095 | -0.022 | 0.014 | 0.119 | -0.049 | 0.006 | 386 |
| | | Q4 | -0.133 | -0.032 | 0.015 | 0.028 | -0.061 | -0.004 | 386 |

Note: St. = Standardized; SE = Standard Error; p = p-value; lowCI = lower 95% confidence interval; upCI = upper 95% confidence interval.

Supplementary Table 5: Linear regression analysis of epigenetic age estimates calculated by all available six epigenetic clocks on quartiles of Selenoprotein P in the subgroup of men. Model 1 is unadjusted. Model 2 is adjusted for chronological age, sex, BMI, smoking (packyears), and the first four genetic principal components (PC1 to PC4). The first quartile is used as reference.

| Clock | Model | | St. $\beta$ | $\beta$ | SE | p | lowCI | upCI | n |
|---|---|---|---|---|---|---|---|---|---|
| 7-CpG DNAmAA | 1 | Q2 | -0.054 | -0.865 | 0.811 | 0.286 | -2.458 | 0.727 | 580 |
| | | Q3 | -0.074 | -1.202 | 0.818 | 0.142 | -2.809 | 0.405 | 580 |
| | | Q4 | -0.062 | -1.002 | 0.812 | 0.218 | -2.597 | 0.594 | 580 |
| | 2 | Q2 | -0.017 | -0.261 | 1.097 | 0.812 | -2.418 | 1.897 | 326 |
| | | Q3 | -0.033 | -0.527 | 1.137 | 0.643 | -2.764 | 1.709 | 326 |
| | | Q4 | 0.010 | 0.153 | 1.111 | 0.890 | -2.033 | 2.340 | 326 |
| Horvath DNAmAA | 1 | Q2 | -0.029 | -0.272 | 0.623 | 0.663 | -1.498 | 0.954 | 374 |
| | | Q3 | -0.082 | -0.829 | 0.652 | 0.204 | -2.110 | 0.452 | 374 |
| | | Q4 | -0.050 | -0.482 | 0.635 | 0.449 | -1.730 | 0.767 | 374 |
| | 2 | Q2 | 0.011 | 0.102 | 0.677 | 0.880 | -1.230 | 1.433 | 322 |
| | | Q3 | -0.037 | -0.358 | 0.701 | 0.610 | -1.736 | 1.021 | 322 |
| | | Q4 | -0.033 | -0.312 | 0.686 | 0.649 | -1.663 | 1.038 | 322 |
| Hannum DNAmAA | 1 | Q2 | 0.093 | 0.730 | 0.513 | 0.156 | -0.279 | 1.739 | 374 |
| | | Q3 | -0.072 | -0.601 | 0.536 | 0.263 | -1.656 | 0.453 | 374 |
| | | Q4 | -0.027 | -0.216 | 0.522 | 0.679 | -1.244 | 0.811 | 374 |
| | 2 | Q2 | 0.113 | 0.882 | 0.571 | 0.124 | -0.242 | 2.007 | 322 |
| | | Q3 | -0.023 | -0.192 | 0.592 | 0.746 | -1.356 | 0.972 | 322 |
| | | Q4 | -0.018 | -0.146 | 0.579 | 0.802 | -1.286 | 0.994 | 322 |
| PhenoAge DNAmAA | 1 | Q2 | -0.028 | -0.288 | 0.675 | 0.670 | -1.616 | 1.040 | 374 |
| | | Q3 | -0.050 | -0.544 | 0.706 | 0.441 | -1.932 | 0.844 | 374 |
| | | Q4 | -0.078 | -0.820 | 0.688 | 0.234 | -2.172 | 0.532 | 374 |
| | 2 | Q2 | -0.005 | -0.046 | 0.733 | 0.950 | -1.488 | 1.395 | 322 |
| | | Q3 | -0.058 | -0.611 | 0.758 | 0.421 | -2.103 | 0.881 | 322 |
| | | Q4 | -0.100 | -1.039 | 0.743 | 0.163 | -2.501 | 0.422 | 322 |
| GrimAge DNAmAA | 1 | Q2 | 0.020 | 0.134 | 0.441 | 0.761 | -0.733 | 1.001 | 374 |
| | | Q3 | -0.067 | -0.482 | 0.461 | 0.297 | -1.388 | 0.425 | 374 |
| | | Q4 | -0.012 | -0.082 | 0.449 | 0.855 | -0.965 | 0.801 | 374 |
| | 2 | Q2 | 0.031 | 0.209 | 0.446 | 0.640 | -0.669 | 1.087 | 322 |
| | | Q3 | -0.024 | -0.166 | 0.462 | 0.719 | -1.075 | 0.743 | 322 |
| | | Q4 | 0.031 | 0.215 | 0.453 | 0.636 | -0.676 | 1.105 | 322 |
| DunedinPACE | 1 | Q2 | -0.042 | -0.011 | 0.015 | 0.489 | -0.041 | 0.019 | 410 |
| | | Q3 | -0.055 | -0.015 | 0.016 | 0.361 | -0.046 | 0.017 | 410 |
| | | Q4 | -0.102 | -0.025 | 0.015 | 0.098 | -0.055 | 0.005 | 410 |
| | 2 | Q2 | -0.048 | -0.012 | 0.017 | 0.463 | -0.045 | 0.021 | 354 |
| | | Q3 | -0.036 | -0.010 | 0.017 | 0.578 | -0.044 | 0.024 | 354 |
| | | Q4 | -0.079 | -0.020 | 0.017 | 0.222 | -0.053 | 0.012 | 354 |

Note: St. = Standardized; SE = Standard Error; p = p-value; lowCI = lower 95% confidence interval; upCI = upper 95% confidence interval.

Supplementary Table 6: Linear regression analysis of epigenetic age estimates calculated by all available six epigenetic clocks on quartiles of GPx3 intensity in men and women. Model 1 is unadjusted. Model 2 is adjusted for chronological age, sex, BMI, smoking (packyears), and the first four genetic principal components (PC1 to PC4). The first quartile is used as reference.

| Clock | Model | | St. $\beta$ | $\beta$ | SE | p | lowCI | upCI | n |
|---|---|---|---|---|---|---|---|---|---|
| 7-CpG DNAmAA | 1 | Q2 | -0,027 | -0,432 | 0,594 | 0,467 | -1,598 | 0,734 | 1066 |
| | | Q3 | -0,024 | -0,390 | 0,595 | 0,513 | -1,558 | 0,778 | 1066 |
| | | Q4 | -0,040 | -0,634 | 0,593 | 0,285 | -1,797 | 0,528 | 1066 |
| | 2 | Q2 | 0,012 | 0,187 | 0,791 | 0,813 | -1,365 | 1,740 | 614 |
| | | Q3 | 0,008 | 0,127 | 0,805 | 0,875 | -1,453 | 1,707 | 614 |
| | | Q4 | -0,042 | -0,670 | 0,801 | 0,403 | -2,243 | 0,903 | 614 |
| Horvath DNAmAA | 1 | Q2 | -0,033 | -0,322 | 0,462 | 0,487 | -1,228 | 0,585 | 689 |
| | | Q3 | -0,076 | -0,772 | 0,471 | 0,102 | -1,697 | 0,153 | 689 |
| | | Q4 | -0,064 | -0,628 | 0,458 | 0,171 | -1,527 | 0,272 | 689 |
| | 2 | Q2 | -0,016 | -0,151 | 0,478 | 0,753 | -1,090 | 0,788 | 608 |
| | | Q3 | -0,048 | -0,475 | 0,489 | 0,332 | -1,436 | 0,486 | 608 |
| | | Q4 | -0,030 | -0,284 | 0,485 | 0,558 | -1,236 | 0,667 | 608 |
| Hannum DNAmAA | 1 | Q2 | 0,009 | 0,072 | 0,377 | 0,848 | -0,668 | 0,813 | 689 |
| | | Q3 | -0,017 | -0,140 | 0,385 | 0,716 | -0,895 | 0,615 | 689 |
| | | Q4 | -0,059 | -0,472 | 0,374 | 0,208 | -1,206 | 0,263 | 689 |
| | 2 | Q2 | 0,007 | 0,055 | 0,390 | 0,887 | -0,712 | 0,822 | 608 |
| | | Q3 | 0,002 | 0,017 | 0,399 | 0,967 | -0,768 | 0,801 | 608 |
| | | Q4 | -0,021 | -0,166 | 0,396 | 0,675 | -0,943 | 0,611 | 608 |
| PhenoAge DNAmAA | 1 | Q2 | -0,034 | -0,365 | 0,498 | 0,464 | -1,343 | 0,613 | 689 |
| | | Q3 | -0,090 | -0,989 | 0,508 | 0,052 | -1,986 | 0,008 | 689 |
| | | Q4 | -0,098 | -1,028 | 0,494 | 0,038 | -1,998 | -0,059 | 689 |
| | 2 | Q2 | -0,031 | -0,324 | 0,521 | 0,534 | -1,346 | 0,699 | 608 |
| | | Q3 | -0,102 | -1,087 | 0,532 | 0,042 | -2,133 | -0,042 | 608 |
| | | Q4 | -0,061 | -0,629 | 0,527 | 0,233 | -1,665 | 0,406 | 608 |
| GrimAge DNAmAA | 1 | Q2 | 0,004 | 0,026 | 0,332 | 0,937 | -0,625 | 0,677 | 689 |
| | | Q3 | -0,059 | -0,430 | 0,338 | 0,203 | -1,094 | 0,233 | 689 |
| | | Q4 | -0,176 | -1,246 | 0,329 | 0,000 | -1,892 | -0,601 | 689 |
| | 2 | Q2 | -0,044 | -0,313 | 0,310 | 0,313 | -0,923 | 0,296 | 608 |
| | | Q3 | -0,085 | -0,628 | 0,318 | 0,048 | -1,252 | -0,005 | 608 |
| | | Q4 | -0,136 | -0,975 | 0,315 | 0,002 | -1,593 | -0,358 | 608 |
| DunedinPACE | 1 | Q2 | -0,006 | -0,002 | 0,011 | 0,885 | -0,023 | 0,020 | 764 |
| | | Q3 | -0,044 | -0,011 | 0,011 | 0,309 | -0,033 | 0,010 | 764 |
| | | Q4 | -0,218 | -0,054 | 0,011 | 0,000 | -0,076 | -0,033 | 764 |
| | 2 | Q2 | -0,011 | -0,003 | 0,011 | 0,801 | -0,024 | 0,019 | 674 |
| | | Q3 | -0,039 | -0,010 | 0,011 | 0,369 | -0,032 | 0,012 | 674 |
| | | Q4 | -0,146 | -0,037 | 0,011 | 0,001 | -0,059 | -0,015 | 674 |

Note: St. = Standardized; SE = Standard Error; p = p-value; lowCI = lower 95% confidence interval; upCI = upper 95% confidence interval.

Supplementary Table 7: Linear regression analysis of epigenetic age estimates calculated by all available six epigenetic clocks on quartiles of GPx3 intensity in the subgroup of women. Model 1 is unadjusted. Model 2 is adjusted for chronological age, sex, BMI, smoking (packyears), and the first four genetic principal components (PC1 to PC4). The first quartile is used as reference.

| Clock | Model | | St. $\beta$ | $\beta$ | SE | p | lowCI | upCI | n |
|---|---|---|---|---|---|---|---|---|---|
| 7-CpG DNAmAA | 1 | Q2 | -0,115 | -1,804 | 0,832 | 0,031 | -3,439 | -0,170 | 548 |
| | | Q3 | -0,076 | -1,159 | 0,814 | 0,155 | -2,758 | 0,440 | 548 |
| | | Q4 | -0,043 | -0,638 | 0,793 | 0,421 | -2,195 | 0,919 | 548 |
| | 2 | Q2 | -0,052 | -0,840 | 1,124 | 0,455 | -3,051 | 1,372 | 324 |
| | | Q3 | 0,012 | 0,198 | 1,136 | 0,862 | -2,038 | 2,433 | 324 |
| | | Q4 | -0,028 | -0,406 | 1,058 | 0,701 | -2,487 | 1,675 | 324 |
| Horvath DNAmAA | 1 | Q2 | -0,012 | -0,120 | 0,650 | 0,853 | -1,398 | 1,157 | 362 |
| | | Q3 | -0,024 | -0,242 | 0,666 | 0,717 | -1,552 | 1,068 | 362 |
| | | Q4 | 0,016 | 0,144 | 0,604 | 0,812 | -1,044 | 1,332 | 362 |
| | 2 | Q2 | 0,007 | 0,074 | 0,693 | 0,915 | -1,289 | 1,438 | 321 |
| | | Q3 | 0,016 | 0,165 | 0,708 | 0,816 | -1,229 | 1,559 | 321 |
| | | Q4 | 0,055 | 0,493 | 0,652 | 0,451 | -0,791 | 1,776 | 321 |
| Hannum DNAmAA | 1 | Q2 | 0,019 | 0,141 | 0,493 | 0,774 | -0,828 | 1,111 | 362 |
| | | Q3 | 0,002 | 0,016 | 0,505 | 0,974 | -0,977 | 1,010 | 362 |
| | | Q4 | -0,025 | -0,169 | 0,458 | 0,713 | -1,070 | 0,733 | 362 |
| | 2 | Q2 | 0,061 | 0,452 | 0,521 | 0,387 | -0,574 | 1,477 | 321 |
| | | Q3 | 0,058 | 0,448 | 0,533 | 0,401 | -0,600 | 1,496 | 321 |
| | | Q4 | 0,009 | 0,058 | 0,491 | 0,906 | -0,907 | 1,023 | 321 |
| PhenoAge DNAmAA | 1 | Q2 | -0,032 | -0,348 | 0,711 | 0,625 | -1,746 | 1,051 | 362 |
| | | Q3 | -0,039 | -0,436 | 0,729 | 0,550 | -1,869 | 0,998 | 362 |
| | | Q4 | -0,069 | -0,676 | 0,661 | 0,307 | -1,976 | 0,624 | 362 |
| | 2 | Q2 | -0,048 | -0,519 | 0,751 | 0,490 | -1,998 | 0,959 | 321 |
| | | Q3 | -0,072 | -0,801 | 0,768 | 0,298 | -2,312 | 0,711 | 321 |
| | | Q4 | -0,043 | -0,417 | 0,707 | 0,555 | -1,809 | 0,974 | 321 |
| GrimAge DNAmAA | 1 | Q2 | -0,016 | -0,106 | 0,426 | 0,804 | -0,943 | 0,731 | 362 |
| | | Q3 | -0,017 | -0,112 | 0,436 | 0,797 | -0,971 | 0,746 | 362 |
| | | Q4 | -0,131 | -0,777 | 0,396 | 0,050 | -1,555 | 0,002 | 362 |
| | 2 | Q2 | -0,066 | -0,439 | 0,437 | 0,316 | -1,300 | 0,421 | 321 |
| | | Q3 | -0,068 | -0,469 | 0,447 | 0,295 | -1,348 | 0,411 | 321 |
| | | Q4 | -0,135 | -0,815 | 0,412 | 0,049 | -1,625 | -0,005 | 321 |
| DunedinPACE | 1 | Q2 | -0,095 | -0,022 | 0,014 | 0,114 | -0,050 | 0,005 | 407 |
| | | Q3 | -0,073 | -0,018 | 0,014 | 0,221 | -0,046 | 0,011 | 407 |
| | | Q4 | -0,308 | -0,067 | 0,013 | 0,000 | -0,093 | -0,041 | 407 |
| | 2 | Q2 | -0,096 | -0,023 | 0,014 | 0,113 | -0,051 | 0,005 | 361 |
| | | Q3 | -0,089 | -0,021 | 0,014 | 0,140 | -0,050 | 0,007 | 361 |
| | | Q4 | -0,234 | -0,051 | 0,014 | 0,000 | -0,078 | -0,024 | 361 |

Note: St. = Standardized; SE = Standard Error; p = p-value; lowCI = lower 95% confidence interval; upCI = upper 95% confidence interval.

Supplementary Table 8: Linear regression analysis of epigenetic age estimates calculated by all available six epigenetic clocks on quartiles of GPx3 intensity in the subgroup of men. Model 1 is unadjusted. Model 2 is adjusted for chronological age, sex, BMI, smoking (packyears), and the first four genetic principal components (PC1 to PC4). The first quartile is used as reference.

| Clock | Model | | St. $\beta$ | $\beta$ | SE | p | lowCI | upCI | n |
|---|---|---|---|---|---|---|---|---|---|
| 7-CpG DNAmAA | 1 | Q2 | 0,055 | 0,855 | 0,826 | 0,301 | -0,767 | 2,478 | 518 |
| | | Q3 | 0,037 | 0,608 | 0,849 | 0,474 | -1,060 | 2,277 | 518 |
| | | Q4 | -0,020 | -0,343 | 0,874 | 0,695 | -2,059 | 1,373 | 518 |
| | 2 | Q2 | 0,064 | 0,992 | 1,124 | 0,378 | -1,221 | 3,205 | 290 |
| | | Q3 | -0,014 | -0,221 | 1,160 | 0,849 | -2,504 | 2,063 | 290 |
| | | Q4 | -0,068 | -1,196 | 1,248 | 0,338 | -3,652 | 1,260 | 290 |
| Horvath DNAmAA | 1 | Q2 | -0,054 | -0,510 | 0,635 | 0,423 | -1,760 | 0,740 | 327 |
| | | Q3 | -0,134 | -1,296 | 0,645 | 0,045 | -2,564 | -0,028 | 327 |
| | | Q4 | -0,093 | -0,999 | 0,696 | 0,152 | -2,369 | 0,371 | 327 |
| | 2 | Q2 | -0,036 | -0,324 | 0,667 | 0,628 | -1,637 | 0,990 | 287 |
| | | Q3 | -0,121 | -1,127 | 0,688 | 0,103 | -2,482 | 0,227 | 287 |
| | | Q4 | -0,124 | -1,296 | 0,741 | 0,082 | -2,756 | 0,163 | 287 |
| Hannum DNAmAA | 1 | Q2 | 0,001 | 0,008 | 0,546 | 0,988 | -1,065 | 1,081 | 327 |
| | | Q3 | -0,038 | -0,317 | 0,554 | 0,567 | -1,406 | 0,772 | 327 |
| | | Q4 | -0,021 | -0,190 | 0,598 | 0,750 | -1,367 | 0,986 | 327 |
| | 2 | Q2 | -0,037 | -0,299 | 0,594 | 0,615 | -1,467 | 0,870 | 287 |
| | | Q3 | -0,047 | -0,385 | 0,612 | 0,530 | -1,591 | 0,820 | 287 |
| | | Q4 | -0,040 | -0,366 | 0,659 | 0,580 | -1,664 | 0,932 | 287 |
| PhenoAge DNAmAA | 1 | Q2 | -0,037 | -0,381 | 0,694 | 0,584 | -1,746 | 0,985 | 327 |
| | | Q3 | -0,143 | -1,514 | 0,704 | 0,032 | -2,899 | -0,128 | 327 |
| | | Q4 | -0,094 | -1,101 | 0,761 | 0,149 | -2,598 | 0,395 | 327 |
| | 2 | Q2 | -0,010 | -0,095 | 0,729 | 0,896 | -1,531 | 1,341 | 287 |
| | | Q3 | -0,145 | -1,489 | 0,752 | 0,049 | -2,970 | -0,007 | 287 |
| | | Q4 | -0,076 | -0,873 | 0,810 | 0,282 | -2,469 | 0,722 | 287 |
| GrimAge DNAmAA | 1 | Q2 | 0,022 | 0,152 | 0,457 | 0,740 | -0,747 | 1,051 | 327 |
| | | Q3 | -0,109 | -0,763 | 0,464 | 0,101 | -1,675 | 0,149 | 327 |
| | | Q4 | -0,137 | -1,060 | 0,501 | 0,035 | -2,045 | -0,075 | 327 |
| | 2 | Q2 | -0,035 | -0,240 | 0,447 | 0,592 | -1,121 | 0,640 | 287 |
| | | Q3 | -0,126 | -0,873 | 0,461 | 0,059 | -1,781 | 0,035 | 287 |
| | | Q4 | -0,147 | -1,146 | 0,497 | 0,022 | -2,124 | -0,168 | 287 |
| DunedinPACE | 1 | Q2 | 0,080 | 0,020 | 0,016 | 0,209 | -0,011 | 0,051 | 357 |
| | | Q3 | -0,015 | -0,004 | 0,016 | 0,809 | -0,036 | 0,028 | 357 |
| | | Q4 | -0,080 | -0,022 | 0,017 | 0,200 | -0,056 | 0,012 | 357 |
| | 2 | Q2 | 0,064 | 0,016 | 0,017 | 0,345 | -0,017 | 0,049 | 313 |
| | | Q3 | 0,006 | 0,001 | 0,017 | 0,934 | -0,033 | 0,036 | 313 |
| | | Q4 | -0,061 | -0,017 | 0,018 | 0,354 | -0,053 | 0,019 | 313 |

Note: St. = Standardized; SE = Standard Error; p = p-value; lowCI = lower 95% confidence interval; upCI = upper 95% confidence interval.

Figures

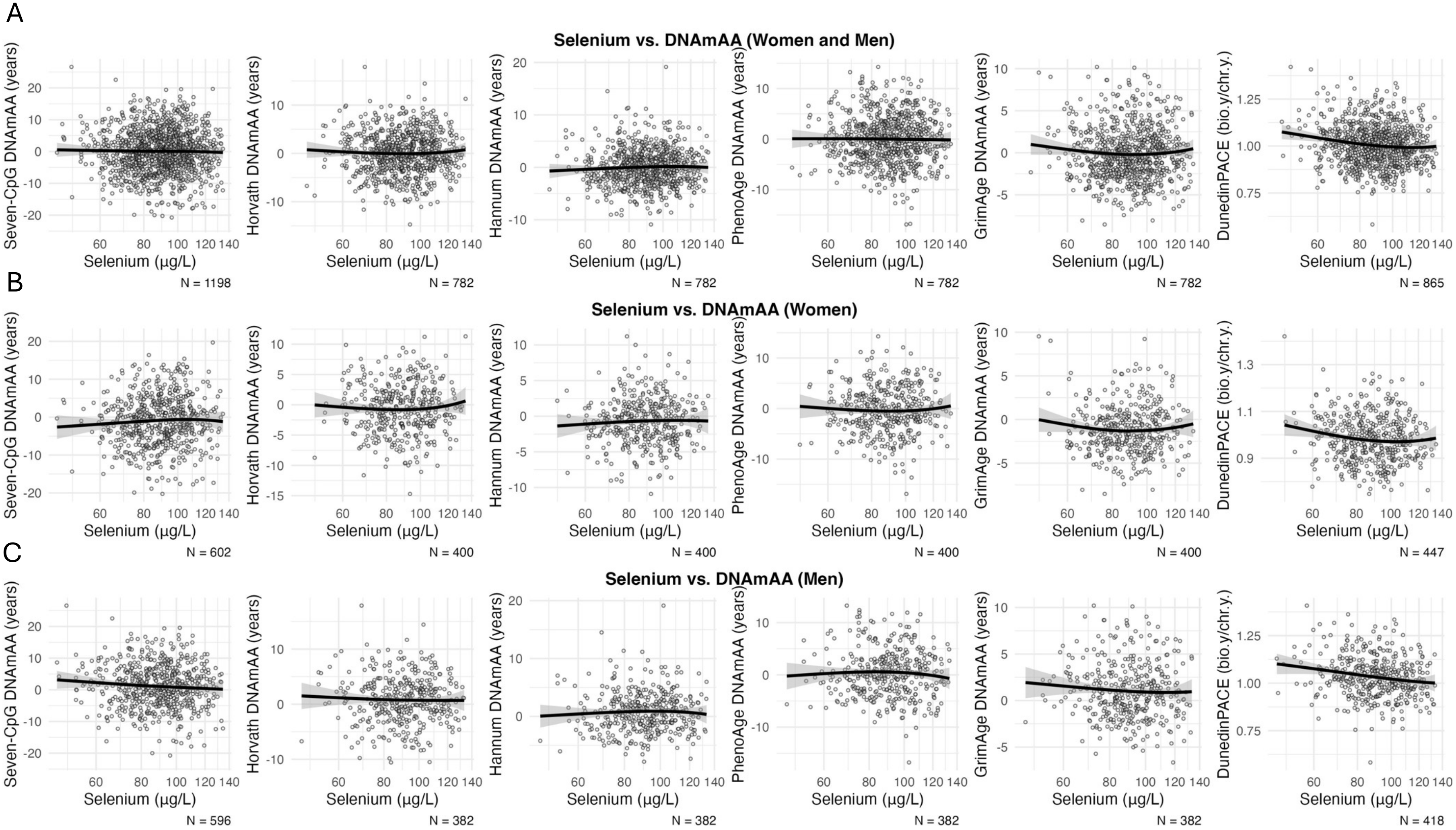


Supplementary Figure 1: Scatterplots of serum selenium levels and biological age estimators calculated from all six available epigenetic clocks in women and men (A) as well as in the all-women (B) and all-me (C) subgroup. The x-axis is log-scaled. All available participants of the older age group are included.

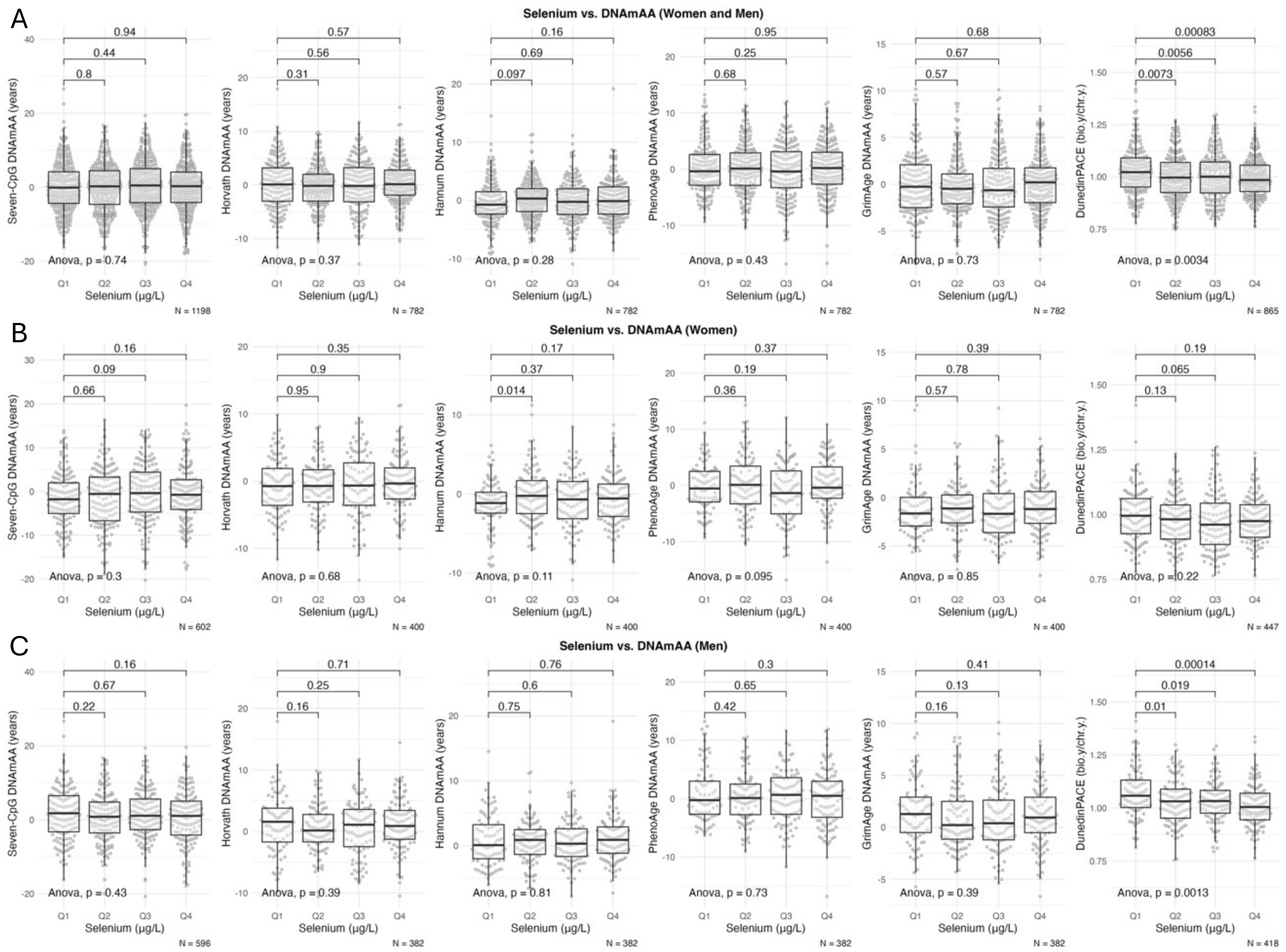


Supplementary Table 2: Boxplots of biological age estimators calculated by all six available epigenetic clocks stratified serum selenium status (deficient vs. sufficient) in men and women (A), women (B) and men (C). Statistical significance of difference between group means was assessed by t-test.

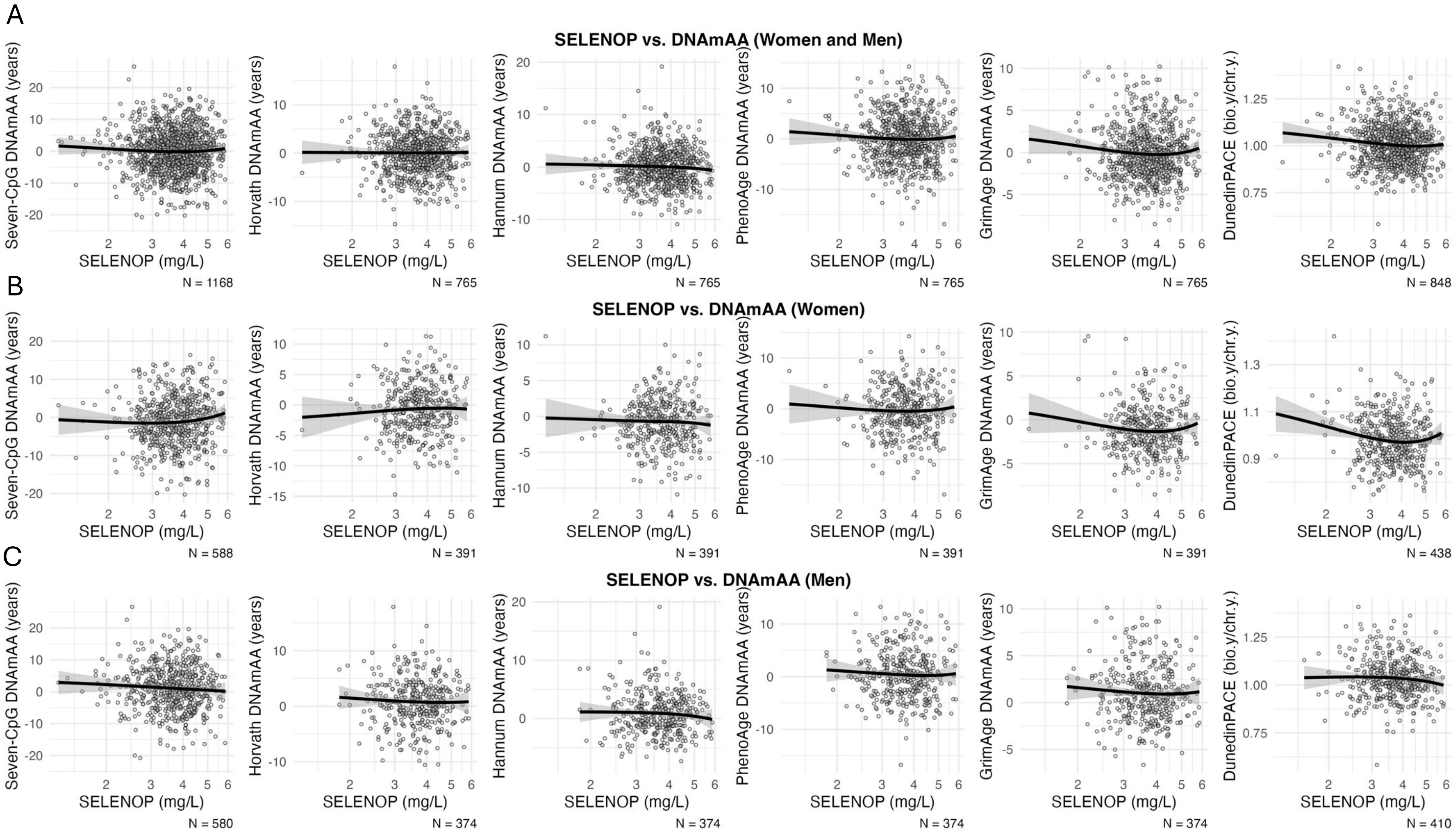


Supplementary Figure 3: Scatterplots of selenoprotein P levels and biologcal age estimators calculated from all six available epigenetic clocks in women and men (A) as well as in the all-women (B) and all-me (C) subgroup. The x-axis is log-scaled. All available participants of the older age group are included.

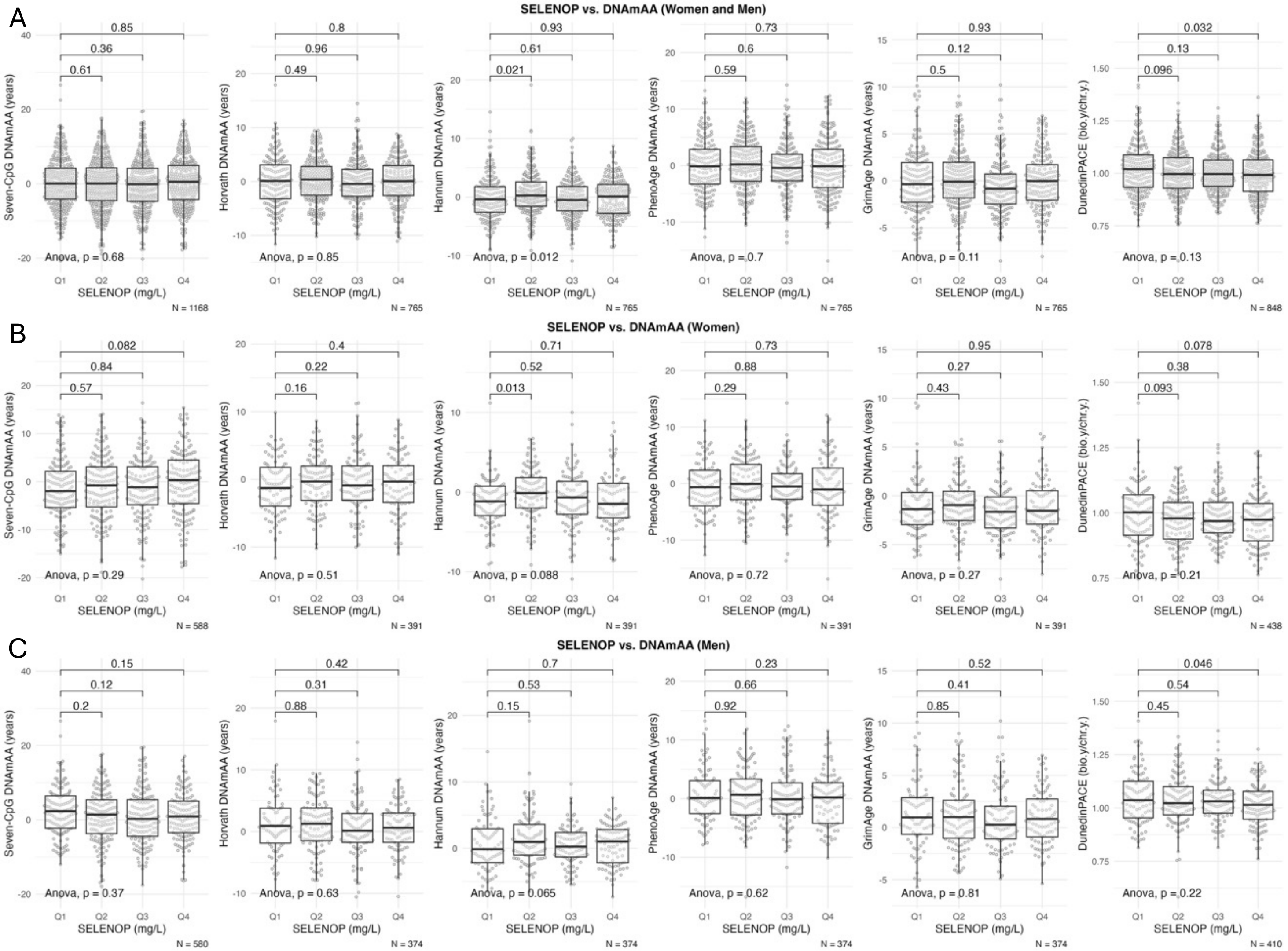


Supplementary Figure 4: Boxplots of biological age estimators calculated by all six available epigenetic clocks stratified by quartiles of selenoprotein P in men and women (A), women (B) and men (C). Statistical significance of difference between group means was assessed by t-test.

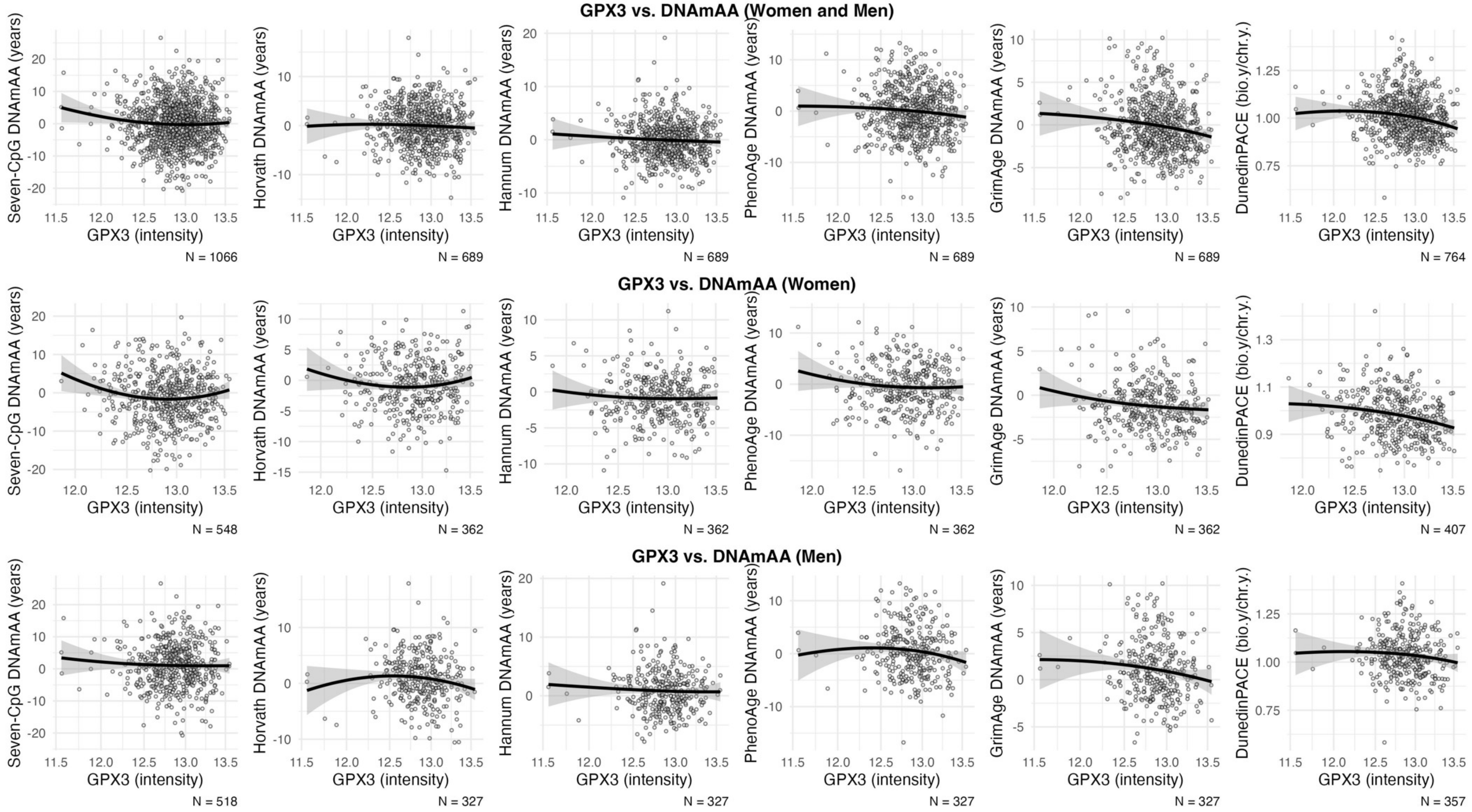


Supplementary Figure 3: Scatterplots of GPx3 intensity and biological age estimators calculated from all six available epigenetic clocks in women and men (A) as well as in the all-women (B) and all-me (C) subgroup. The x-axis is log-scaled. All available participants of the older age group are included.

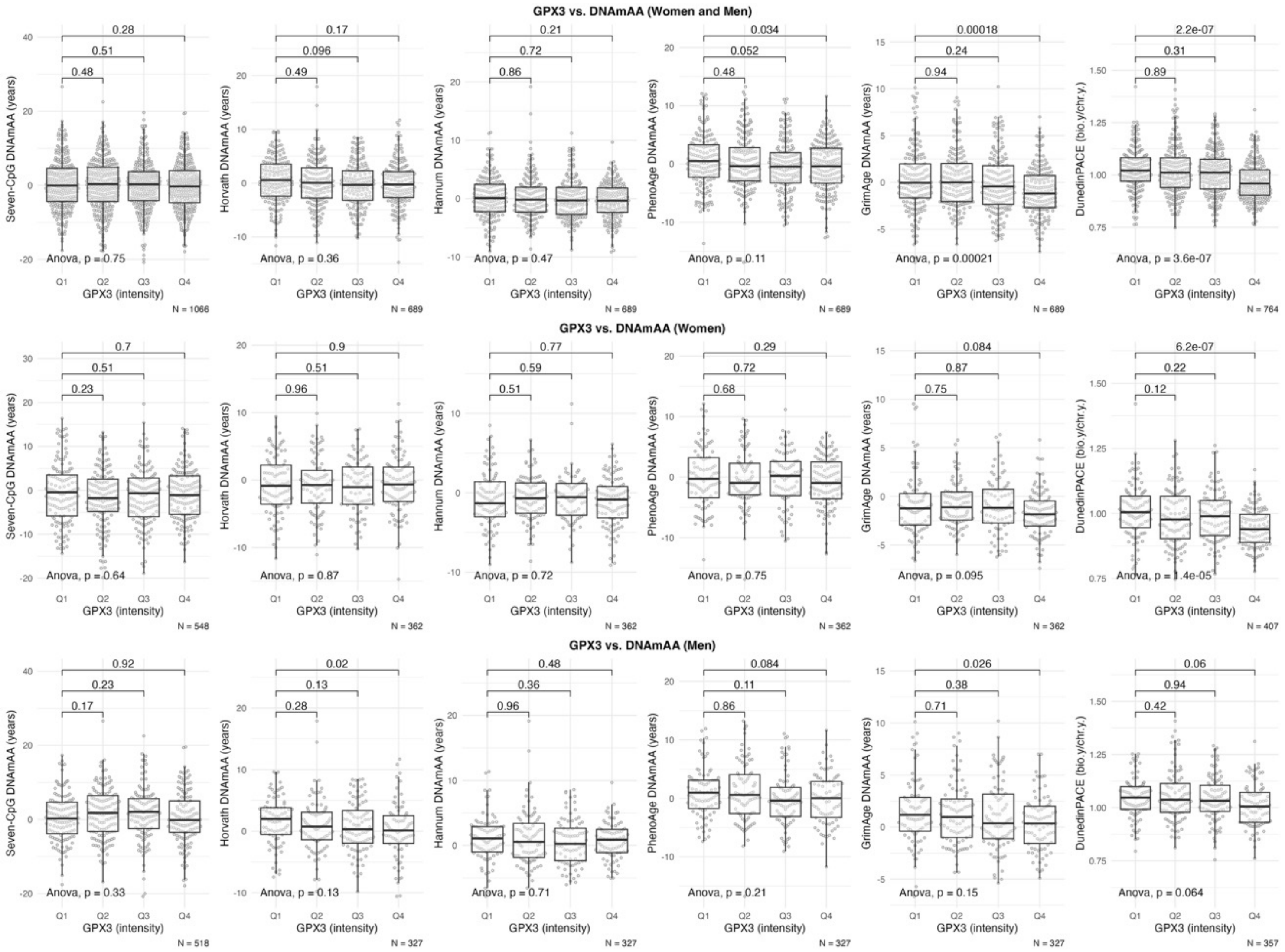


Supplementary Table 4: Boxplots of biological age estimators calculated by all six available epigenetic clocks stratified by quartiles of GPx3 intensities (proteomics) in men and women (A), women (B) and men (C). Statistical significance of difference between group means was assessed by t-test.
